# The fecal microbiome in diverticulitis and asymptomatic diverticulosis: A case-control study in the US

**DOI:** 10.1101/19001404

**Authors:** Meredith A. J. Hullar, Richard S. Sandstrom, John A. Stamatoyannopoulos, Johanna W. Lampe, Lisa L. Strate

## Abstract

**Objective:** The intestinal microbiota are hypothesized to play a role in the pathogenesis of diverticulitis. We compared fecal microbial communities in individuals with diverticulitis to those with uncomplicated diverticulosis.

**Methods:** We used 16S ribosomal RNA gene sequencing to assess and compare the microbiota composition of fecal samples from 10 patients presenting with acute diverticulitis (cases) and 10 controls with asymptomatic diverticulosis matched on age and sex.

**Results:** We found differences in the distribution of relative abundances of bacterial phyla and genera in diverticulitis cases versus diverticulosis controls. At the phyla level, Verrucomicrobia was more abundant on average in diverticulitis cases at the time of diagnosis than in diverticulosis controls (p=0.07). Univariate analysis identified a significant increase in the genera Coriobacteria (p=0.050), Anaerotruncus (0.046), Subdoliganulum (p=0.034), Marvinabryantia (p=0.006), and Akkermansia (p=0.04), and a decrease in Barnesiella (p=0.035) and Coprococcus (p=0.035) in diverticulitis cases at the time of diagnosis compared to diverticulosis controls. However, after correction for multiple comparisons, these differences were no longer significant. Partial least squares discriminant analysis on all microbial genera showed partial separation of diverticulitis cases at diagnosis and diverticulosis controls. The microbial alpha diversity was higher in diverticulitis cases at time of diagnosis vs controls but this was not significant (Shannon diversity index 7.4±0.6 vs 6.8±0.7, p=0.08).

**Conclusions:** Individuals with diverticulitis differ from those with asymptomatic diverticulosis based on components of the fecal microbiome.

## Introduction

Diverticulosis is one of the most common gastrointestinal disorders in Western societies.(1-3) In the US, more than $2 billion are spent annually on hospitalizations for diverticulitis.(1) This figure does not account for the costs associated with the 3 million outpatient visits for diverticulitis each year, or for the indirect costs associated with this disease.(1)

The etiopathogenesis of diverticulitis remains unknown. Traditional theories postulate that diverticulitis occurs when a diverticular sac is obstructed by a fecalith or food particle resulting in trauma, local ischemia, and bacterial stasis.(4) However, there are no data to support this theory, and the importance of the intestinal microbiota in other intestinal disorders has inspired revised disease concepts. Newer theories posit that alterations in the intestinal microbiota composition in patients with diverticulosis lead to chronic mucosal inflammation and ultimately to acute diverticulitis.(5-7) Indirect evidence supports this theory. The pathophysiology of diverticulitis involves the translocation of bacteria across the colon mucosal barrier and standard treatment includes antibiotics.(8) Well-established risk factor for diverticulitis, including dietary red meat,(9) dietary fiber,(10-12) and obesity,(13, 14) influence the gut microbiota composition and function.(15-21) Global geographical differences in the intestinal microbial community also correspond to differences in the prevalence of diverticular disease.(21-23) Several studies have examined the intestinal microbial composition in patients with symptomatic asymptomatic diverticular disease (24-26) and in asymptomatic diverticulosis.(27, 28) Only two small studies have explored the intestinal microbiota in patients with diverticulitis.(29, 30)

Currently, we lack methods to identify individuals with diverticulosis who are at high risk of diverticulitis. This is important because more than half of adults in developed countries have diverticulosis, but fewer than 5% of these individuals will experience diverticulitis.(31, 32) Furthermore, there are no proven means to prevent incident or recurrent diverticulitis short of prophylactic colectomy.(33)

A better understanding of the intestinal microbiota composition in patients with diverticulitis as compared to patients with asymptomatic diverticulosis could lead to the development of a risk-stratification tool as well as preventive interventions. Therefore, we profiled and compared the fecal microbiota in individuals with acute diverticulitis and individuals with asymptomatic diverticulosis using high-resolution, next-generation sequencing of 16S ribosomal RNA (rRNA) gene to assess whether specific differences in the microbial communities could be identified.

## Methods

### Study Participants

The study population included 10 patients with diverticulitis and 10 controls with asymptomatic diverticulosis from the University of Washington Medical Center, a quaternary care center, and Harborview Medical Center, a trauma and safety net hospital in Seattle, WA. Cases of acute diverticulitis were identified and recruited at the time of presentation, and the first stool after diagnosis was sampled. However, all cases were treated promptly and received antibiotics prior to stool collection (range 1 to 57 hours). All diagnoses of diverticulitis were confirmed by abdominal CT scanning. Diverticulitis cases were excluded if emergency surgery was planned for perforation or peritonitis as these patients usually did not have a bowel movement prior to surgery and post-surgical anatomy may alter the microbiome. Controls with asymptomatic diverticulosis matched on age and sex to cases were identified from colonoscopy reports and contacted regarding interest in study participation. Both cases and controls were excluded if they had a history of bowel resection, gastrointestinal cancer, inflammatory bowel disease, irritable bowel syndrome, ongoing chemotherapy or immunosuppression, or antibiotic therapy or bowel preparation in the previous 3 months (except for antibiotics given for the index diverticulitis episode). All participants provided written informed consent and the study was approved by the institutional review board at the University of Washington School of Medicine.

At a one-time study visit, data were obtained regarding past medical history including gastrointestinal illnesses and symptoms, current medication use and smoking history (in pack years of exposure). In addition, height, weight and waist and hip circumference measurements were obtained in clinic by trained study staff, and body mass index (BMI; kg/m^2^) was calculated. At the time of each stool sample collection, participants also completed a self-administered, validated 120-item food frequency questionnaire (FFQ) developed by the Nutrition Assessment Shared Resource, Fred Hutchinson Cancer Research Center, Seattle, WA.(34)

### Fecal Sample Collection and Processing

Two fecal samples were obtained from each participant, one at the time of diagnosis (diverticulitis cases) or study recruitment (diverticulosis controls) and another at least 45 days later (post-treatment for cases). This sampling schema was used to account for possible changes in the microbiota due to acute inflammation and/or antibiotic exposure. We used methods previously tested by our group for fecal collection, preservation and DNA extraction.(35) Briefly, participants collected fecal samples using a home fecal collection kit. Two pea-sized samples were immediately placed in a tube containing RNAlater, shaken vigorously to disperse the sample, and sent to the lab where they were frozen at −80°C until further processing. Fecal bacterial DNA was extracted using the QIAmp DNA stool minikit (Stool Kit, Qiagen, Valencia, CA).

### 16S Ribosomal RNA Pyrosequencing

The 16S rRNA gene was amplified and sequenced using primers 27f and 519r (V1-V3;)(35) for amplicon pyrosequencing (bTEFAP(36)) at Research and Testing (Shallowater, Texas) using Roche 454 FLX titanium instruments and reagents and following the manufacturer’s guidelines. Sequences have been deposited in the Sequence Read Archive of NCBI under accession number SUB2127005.

### Microbiome Analysis

To classify bacterial taxonomy, sequences were aligned and identified in MOTHUR (V.1.28.0) to the Silva 16S rRNA gene reference alignment (www.arb-silva.de)(37, 38) Sequences were converted to standard FASTA format from .sff files. Sequences were removed if they were <300 bp, had homopolymers >8 bp, more than one mismatch to the forward primer, more than one mismatch to the barcode, or ambiguous bases. Potentially chimeric sequences were removed (39),sequences were denoised (40), and aligned to the Silva 16S rRNA gene reference alignment (www.arb-silva.de) using the NAST algorithm. (41-43) The pre.cluster option in MOTHUR was used to minimize any overestimation errors in microbial diversity.(44) Sequences were grouped into operational taxonomic units (OTU) using the mean Neighbor-Joining algorithm. Sequences that did not align to the appropriate 16S rRNA gene region were removed. Low abundance sequences were merged to the high abundant sequences using the pre.cluster option in MOTHUR to minimize the effect of pyrosequencing errors in overestimating microbial diversity.(<u>32</u>) Bacterial taxa were removed if they represented less than 0.005% of the total sequences. (45, 46) The number of sequences in each genera was converted to the relative percentage of the total sequence abundance per individual for multivariate analysis.

### Statistical Analysis

We computed the Jensen Shannon divergence distances (JSD)(47, 48) on pairs of samples. We then performed a global analysis of the microbiome using non-metric multidimensional scaling (NMS) analysis of the JSD distances at the genera level. Unsupervised clustering using unweighted pair group method with arithmetic mean (UPGMA) was performed followed by multi-response permutation procedures (MRPP) to test for differences in community composition in the clusters.(49) A joint plot was used to show the relationship between the bacterial genera and the ordination scores generated from the JSD matrix. Only the vectors with a correlation of >0.5 relative to the NMS axes were shown. A logistic regression model using the NMS axes that described the microbial community composition, adjusted for time from antibiotic treatment (in days), dietary intake of fat, fiber, animal protein, vegetable protein and total energy, smoking (current, past, never), and BMI was used to test whether global differences in all of the genera in the microbiome could distinguish between participants with diverticulitis and diverticulosis.

Differences in the relative abundance at the phyla and genera level between diverticulitis cases and diverticulosis controls, and pre and post-diverticulitis treatment were tested using the unpaired (Mann-Whitney) or paired non-parametric (Wilcoxon signed-rank) test. We used the Benjamini-Hochberg approach to account for multiple comparisons.(50)

To investigate whether cases and controls segregated based on specific components of the fecal microbial community, we employed partial least squares discriminant analysis (PLSDA). PLSDA was performed to sharpen the separation between predetermined groups of individuals, and is utilized when the number of observations is low and the number of explanatory variable is high.(51) The input variables were arcsine transformed relative abundances of the bacterial genera. To test the predictive performance of the model, we used a leave one out approach. We assessed the consistency between the predicted and actual data using the Q^2^ statistic and considered a value of >0.4 indicative of good discrimination. (52) We used R^2^ to assess the degree of fit to the data along the PLSDA decomposition associated with the x and y axes.(53) To estimate the influence of each bacterial genera on the first two axes of the PLSDA model, we calculated the variable of importance (VIP). VIP values rank input variables according to their ability to distinguish different groups. Values greater than 1 are indicative of variables that have an important contribution to the model. (54) To examine how well this model segregated cases and controls, we tested a 20% random selection of samples and calculated R^2^ and Q^2^ statistics.

We estimated alpha variation (within-person) in the overall microbial composition after rarefaction using the Shannon diversity index.(55) Differences in diversity between diverticulitis cases pre and post-diagnosis and diverticulosis controls were tested using the unpaired or paired Tukey’s t-test.

PC-ORD (version 6.0, Glenenden, OR) and XL-STAT (version 2016.04.3233, New York, NY) were the statistical packages used for the analyses.

## Results

The mean age of study participants was 55 (range 38-67) (Table 1). There were 12 males and 8 females. Most cases of diverticulitis were asymptomatic; 80% modified Hinchey class 1a and 20% Hinchey class 1b (small, pericolic abscess). There were no statistically significant differences between diverticulitis cases and diverticulosis controls in the mean BMI (30.2 vs. 27.7 kg/m^2^, *P*=0.39), mean daily total energy intake (1794 vs. 1520 kcal/day, *P*=0.21), or the proportion of current smokers (20% vs 40%, *P*=0.33). However, on average, cases consumed more fiber than controls (22.4 vs. 15.8 g/day, *P*=0.02). In cases, the time between starting antibiotics for the treatment of diverticulitis and obtaining the first fecal sample ranged from 1 hour to 57 hours (median 22.5 hours). The mean time between finishing antibiotic treatment and obtaining the second fecal sample was 93 days (range 45 to 194). All cases received ciprofloxacin and metronidazole for treatment of diverticulitis. In controls, the mean time between undergoing colonoscopy and the first fecal collection was 263 days (range 99 to 721). One case underwent colonoscopy 465 days prior to the first fecal collection. One diverticulitis case sample was excluded from the pair comparisons due to insufficient fecal material for DNA extraction and analysis.

**Table 1.** Baseline characteristics of the study population.

|  | **Diverticulitis Cases**<br>**(n=10)** | **Matched Controls**<br>**with Diverticulosis**<br>**(n=10)** | ***P* value** |
| --- | --- | --- | --- |
| Age, y, mean (SD) | 54.7 (10.2) | 54.8 (10.3) | 0.98 |
| Male, % | 60 | 60 |  |
| Race, Caucasian, % | 70 | 80 | 0.61 |
| Ethnicity, Hispanic, % | 40 | 0 | 0.01 |
| Smoking, current, % | 20 | 40 | 0.33 |
| Body Mass Index, kg/m<sup>2</sup>, mean (SD) | 30.2 (6.9) | 27.7 (5.3) | 0.39 |
| Total energy, kcal/day, mean (SD) | 1794 (604) | 1520 (324) | 0.11 |
| Dietary fiber, g/day, mean (SD) | 22.4 (10.1) | 15.8 (5.5) | 0.02 |
| Protein, g/day, mean (SD) | 69.0 (26.6) | 63.8 (16.1) | 0.24 |

In total, 7.3 × 10^5^ raw sequences were generated resulting in 14,551 ± 3782 sequences per participant sample after quality control with an average sequence length of 463 base pairs. These sequences represented 58 bacterial genera distributed across 7 phyla (Good’s coverage 0.996±0.003). The predominant phyla were Firmicutes (51%) and Bacteroidetes (42%) (Figure 1). Together, Proteobacteria, Verrucomicrobia, Actinobacteria, Fusobacteria, and Tenericutes represented the remaining 7% of the phyla. We found no significant differences in the microbial communities with respect to the relative abundance at the phyla and genera level between the two fecal samples taken in diverticulosis controls, and therefore, the second samples were removed from the analyses. However, the fecal microbial communities of diverticulitis cases differed at diagnosis and post-treatment. There was an increase in the phyla Bacteroidetes in diverticulitis cases after antibiotic treatment compared to at the time of diagnosis (Figure 1 and Table 2). The genera Coriobacteria, Erysipelotrichaceae, and Akkermansia were significantly reduced in diverticulitis cases after treatment when compared to diverticulitis at diagnosis (Table 3). In addition, we found a significant reduction in microbial diversity in diverticulitis cases after treatment (Shannon diversity index 7.4±0.5 vs 6.5±0.7, *P*=0.03). Therefore, we analyzed the pre and post-treatment fecal samples for diverticulitis cases separately.

**Table 2.** Relative abundance of phyla in fecal samples from diverticulitis cases and diverticulosis controls.

|  | Diverticulitis<br>at diagnosis | Diverticulitis<br>after<br>treatment | Diverticulosis<br>controls | Diverticulitis<br>at diagnosis<br>vs<br>Control<sup>a</sup> | Diverticulitis<br>after<br>treatment vs<br>control<sup>a</sup> | Diverticulitis<br>at diagnosis<br>vs treatment<sup>b</sup> |
| --- | --- | --- | --- | --- | --- | --- |
|  | Mean(SD)<sup>c</sup><br>(n=9)<sup>d</sup> | Mean(SD)<sup>c</sup><br>(n=9) | Mean(SD)<sup>c</sup><br>(n=10) | p-value | p-value | p-value |
| Firmicutes | 55.0(14.9) | 50.0(11.4) | 50.0(16.2) | n.s. | n.s. | n.s. |
| Bacteroidetes | 31.9(13.8) | 45.7(13.4) | 42.9(17.3) | n.s. | n.s. | 0.025<sup>e</sup> |
| Tenericutes | 1.8(2.4) | 0.9(1.6) | 1.0(1.9) | n.s. | n.s. | n.s. |
| Actinobacteria | 3.3(3.5) | 0.8(0.7) | 1.3(1.7) | n.s. | n.s. | n.s. |
| Verrucomicrobia | 3.1(4.3) | 0.2(0.7) | 0.04(0.08) | 0.07 | n.s. | n.s. |
| Proteobacteria | 4.3(7.6) | 1.2(1.2) | 4.6(6.2) | n.s. | n.s. | n.s. |
| Fusobacteria | 0.2(0.5) | 1.2(2.8) | 0.03(0.1) | n.s. | n.s. | n.s. |

n.s., not significant
<sup>a</sup>Mann-Whitney test
<sup>b</sup>Paired, Wilcoxon signed-rank test
<sup>c</sup>Expressed as percentage contribution of phyla to the total phyla in the subject
<sup>d</sup>There are only 9 diverticulitis cases in the paired analysis because one case had only one fecal sample.
<sup>e</sup> Not significant when correcting for multiple comparisons using the Benjamin-Hochberg approach

**Table 3.** Relative abundance of genera in fecal samples from diverticulitis cases and diverticulosis controls.

|  | Diverticulitis<br>at diagnosis | Diverticulitis<br>after<br>treatment | Diverticulosis<br>controls | Diverticulitis<br>at diagnosis<br>vs<br>control<sup>a</sup> | Diverticulitis<br>after<br>treatment vs<br>control<sup>a</sup> | Diverticulitis<br>at diagnosis<br>vs<br>treatment<sup>b</sup> |
| --- | --- | --- | --- | --- | --- | --- |
|  | Mean(SD)<sup>c</sup><br>(n=9)<sup>d</sup> | Mean(SD)<sup>c</sup><br>(n=9) | Mean(SD)<sup>c</sup><br>(n=10) | p-value | p-value | p-value |
| P_Actinobacteria/C_Actinobacteria/<br>O_Coriobacteriales/<br>F_Coriobacteriaceae/<br>G_Coriobacterium | 0.10(0.2) | 0.0 | 0.03(0.07) | 0.050<sup>e</sup> | n.s. | 0.05<sup>e</sup> |
| P_Firmicutes/C_Clostridia/<br>O_Clostridiales/F_Clostridiaceae/<br>G_Anaerotruncus | 0.48(0.4) | 0.37(0.5) | 0.19(0.2) | 0.046<sup>e</sup> | n.s. | n.s. |
| P_Bacteroidetes/C_Bacteroidia/<br>O_Bacteroidales/<br>F_Porphyromonadaceae/<br>G_Barnesiella | 0.40(0.4) | 1.34(2.5) | 1.45(1.5) | 0.035<sup>e</sup> | n.s. | n.s. |
| P_Firmicutes/C_Clostridia/<br>O_Clostridiales/F_Lachnospiraceae/<br>G_Coprococcus | 1.46(1.4) | 2.56(3.2) | 2.33(0.9) | 0.035<sup>e</sup> | n.s. | n.s. |
| P_Firmicutes/<br>C_Clostridia/O_Clostridiales/<br>F_Ruminococcaceae/<br>G_Subdoligranulum | 1.58(1.5) | 0.67(0.7) | 1.44(2.8) | 0.034<sup>e</sup> | n.s. | n.s. |
| P_Firmicutes/C_Clostridia/<br>O_Clostridiales/F_Lachnospiraceae<br>/G_Marvinbryantia | 0.23(0.2) | 0.11(0.1) | 0.05(0.06) | 0.006<sup>e</sup> | n.s. | n.s. |
| Firmicutes/C_Erysipelotrichia/<br>O_Erysipeotrichales/<br>F_Erysipelotrichaceae/ | 0.67(1.4) | 0.04(0.1) | 0.28(0.6) | n.s. | n.s. | 0.03<sup>e</sup> |
| P_Verrucomicrobia/C_Verrucomicrobia/<br>O_Verrucomicrobiales/<br>F_Verrucomicrobiaceae/<br>G_Akkermansia | 3.14(4.0) | 0.19(0.5) | 0.05(0.1) | 0.04<sup>e</sup> | n.s. | 0.02<sup>e</sup> |

n.s., not significant
<sup>a</sup>Mann-Whitney test
<sup>b</sup>Paired, Wilcoxon signed-rank test
<sup>c</sup>Expressed as percentage contribution of phyla to the total phyla in the subject
<sup>d</sup> There are only 9 diverticulitis cases in the paired analysis because one case had only one fecal sample.
<sup>e</sup>Not significant when corrected for multiple comparisons using the Benjamini-Hochberg approach

**Fig 1.**
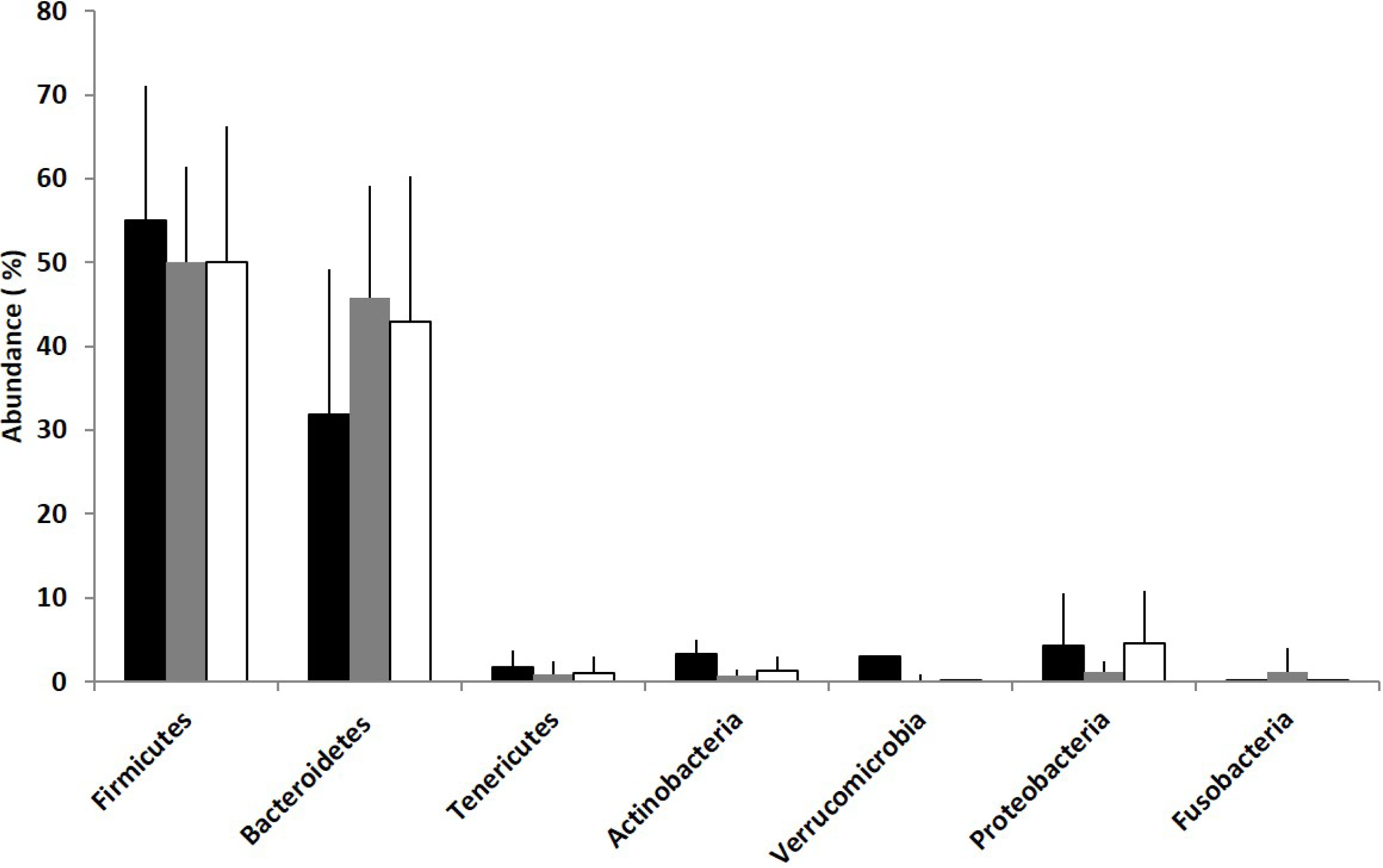
The distribution of bacterial phyla in diverticulitis cases at the time of diagnosis (black bars) and after treatment (dark gray bars) as wells as diverticulosis controls (white bars). There was a borderline significant difference in the abundance of Verrucomicrobia between diverticulitis cases at diagnosis and diverticulosis controls (3.1 vs. 0.04, respectively; p=0.07)

At the phyla level, Verrucomicrobia tended to be more abundant on average in diverticulitis cases at diagnosis than in diverticulosis controls (3.1 vs. 0.04, p=0.07) (Figure 1 and Table 2). We found no significant differences at the phyla level between diverticulitis cases after treatment and diverticulosis controls.

The relative abundances at the genera level are shown in Table 3. Significant differences in Coriobacteria, Anaerotruncus, Barnesiella, Coprococcus, Subdoligranulum, Marvinbryantia, and Akkermansia were seen in diverticulitis cases at diagnosis when compared to diverticulosis controls (Table 3). However, these findings lost significance after adjustment for multiple comparisons. No significant differences were seen in the genera of diverticulitis cases after treatment and diverticulosis controls.

We performed a global analysis of the microbiome at the genera level using multivariate analysis followed by unsupervised clustering. This descriptive analysis tends to cluster participants based on the dominant members of the microbial community.(56) Using nonmetric multidimensional scaling, after 500 iterations and a final stress value=9.35, we determined that 3 axes cumulatively explained 92% of the variation in the fecal microbial community with 64%, 18%, and 13% of the variation in the data attributed to axes 1, 2, and 3, respectively (Supplemental Figure 1). We identified two clusters using an unsupervised clustering approach that were significantly different in their microbiome composition (multiple response permutation procedure Cluster 1 vs. Cluster 2, MRPP; T=-17.96, A=0.24, p<0.0001). Bacteroides (r>0.5) and Ruminococcus (r>0.5) were the most abundant bacterial genera in the first cluster, whereas Prevotella (r>0.5) was the abundant bacterial genera in the second. We included time from antibiotics to fecal sample, diet, smoking and BMI in a logistic regression model. Only time from antibiotic treatment remained significant in the final regression model. After adjustment for time from antibiotic treatment, no significant association was seen between diverticulitis and the overall microbial composition of dominant organisms, as defined via NMS axes.

In a multivariate analysis, using a supervised approach (PLSDA) including all of the bacterial genera, we found that diverticulitis cases at diagnosis tended to separate from diverticulosis controls. (Figure 2). When we randomly added 20% of the samples to the model, we found that we were able to correctly classify samples 79% of the time (Q^2^=0.44 for Axis 1 and 0.43 for Axis 2 and R^2^ =0.10 for Axis 1 and 0.52 for Axis 2). Bacteria with a VIP > 1 and associated with Axis 1 included Alistipes, Faecalibacterium, Coprococcus, and Pseudobutyrivibrio. Akkermansia, Roseburia, Escherichia, Barnesiella, and Subdoligranulum were associated with Axis 2 (VIPs > 1). Genera shared by both Axis 1 and 2 that had VIP >1 included Bacteroides, Ruminococcus, Prevotella, and Blautia. Of these, Bacteroides, Ruminococcus, and Prevotella were associated with the two dominant clusters in the global analysis (see above and Supplemental Figure 1), but as noted above, these genera were not associated with diverticulitis. Akkermansia, Barnesiella, Coprococcus, and Subdolignulum also differed between diverticulitis cases at diagnosis and controls in the univariate analysis (Table 3).

**Fig 2.**
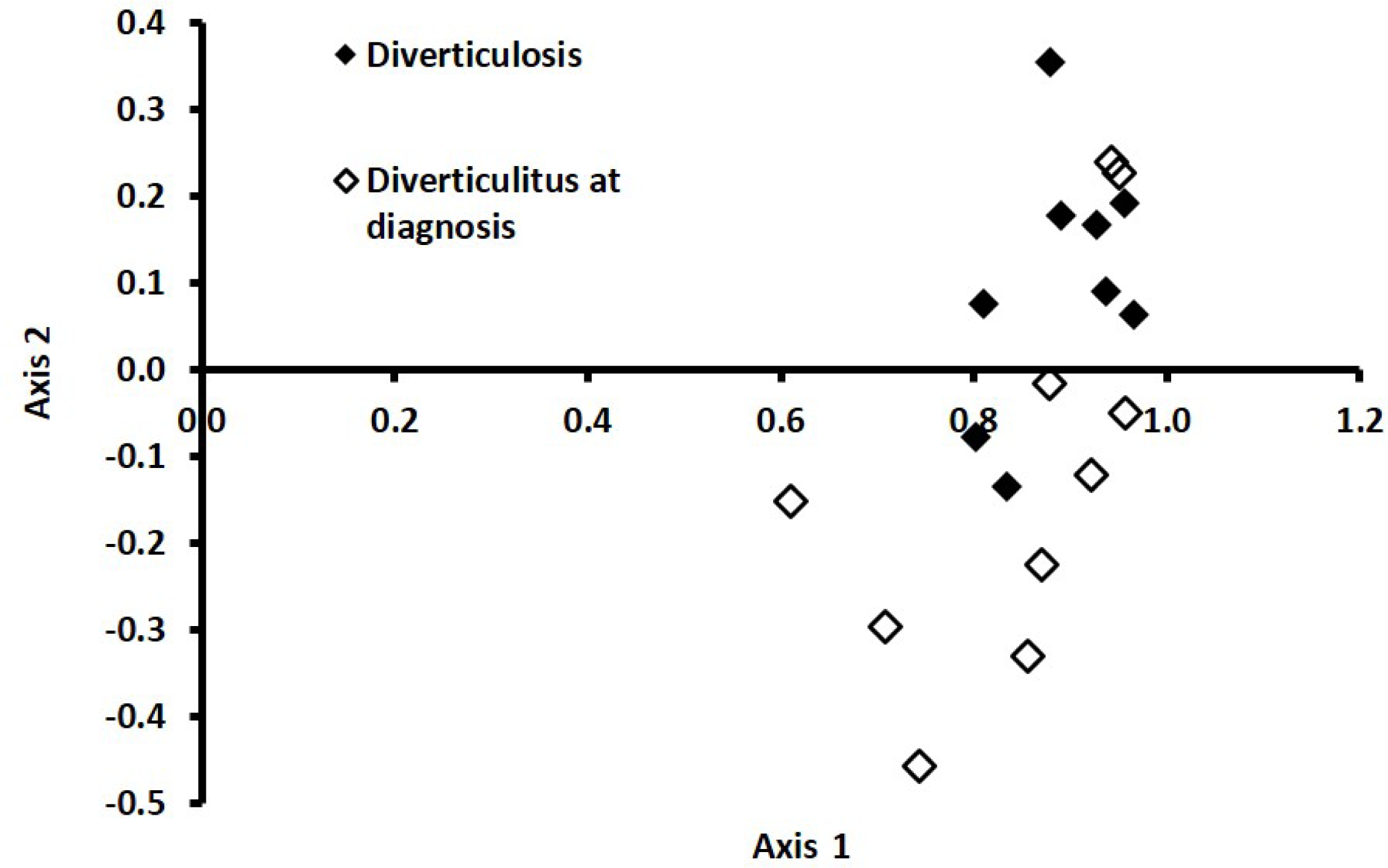
PLSDA of diverticulitis cases at diagnosis (white triangles) and diverticulosis controls (black triangles). Discriminant analysis explained 97% of the variation in the data.

There was a tendency for microbial alpha diversity to be higher in diverticulitis cases at diagnosis vs diverticulosis controls (Shannon diversity index 7.4±0.6 vs 6.8±0.7, p=0.08). The diversity in diverticulitis cases after treatment was not significantly different from controls (Shannon diversity index 6.6±0.7 vs 6.8±0.7, *P*=0.4).

## Discussion

In this study, we found differences in the composition of the fecal microbiota in patients with acute diverticulitis at the time of diagnosis and those with asymptomatic diverticulosis. Differences were seen in the relative abundances of the phyla *Verrucomicrobia*. Significant differences were also seen in the genera Coriobacteria, Anaerotruncus, Barnesiella, Coprococcus, Subdoligranulum, Marvinbryantia, and Akkermansia. The differences at the phyla and genera level lost significance when adjusted for multiple comparisons. Nonetheless, a supervised clustering approach was able to distinguish diverticulitis cases from diverticulosis controls 80% of the time based on the relative abundance of the 7 genera.

A few studies have examined the intestinal microbiota in patients with symptomatic asymptomatic diverticular disease (SUDD).(24, 26) SUDD is an incompletely understood condition generally defined as the presence of abdominal symptoms in patients with diverticulosis and gastrointestinal symptoms in the absence of evidence of diverticulitis. Barbara et al, compared the fecal and mucosal microbiota in 14 controls without diverticulosis, 16 patients with asymptomatic diverticulosis and 8 patients with SUDD.(24) The stool of patients with asymptomatic diverticulosis and SUDD had a lower relative abundance of *Clostridium* cluster IV than controls without diverticulosis, and patients with SUDD had a lower abundance of *Clostridium* cluster IX, *Fusobacterium* and *Lactobacillaceae* compared to those with asymptomatic diverticulosis. Biopsies taken in the area of diverticulosis in patients with SUDD had a lower abundance of *Akkermansia* than in the more proximal colon. In patients with SUDD and asymptomatic diverticulosis, there were lower amounts of *Enterobacteriaceae* at all sites. Tursi and colleagues analyzed the fecal microbiota and metabolome in women with SUDD (n=15), asymptomatic diverticulosis (n=13) and healthy controls (n=16). The amount of *Akkermansia muciniphilia* assessed via quantitative real-time polymerase chain reaction was lower in healthy controls when compared to those with asymptomatic diverticulosis and SUDD. Patients with SUDD had low levels of valerate, butyrate, and choline and high levels of N-acetyl derivatives.(26)

Few studies have examined the intestinal microbiota in patients with diverticulitis. Gueimonde et al compared mucosal-associated *Bifidobacterium* using qualitative and quantitative PCR in 34 patients: diverticulitis (n=9), IBD (n=4), and colon cancer (n=21) and found significantly higher total levels in diverticulitis than in the other diagnoses.(30) Daniels et al studied the microbiota in 31 patients presenting with acute diverticulitis and 25 controls presenting to colonoscopy for a variety of diagnoses including IBD and colon cancer.(29) Microbial material was collected using rectal swabs. PCR amplification of the 16S-23S rDNA interspace region was performed for the phyla Firmicutes, Bacteroidetes, Actinobacteria, Fusobacteria, Verrucomicrobia, and Proteobacteria and species were identified using in silico comparison of fragment lengths. The diversity of Proteobacteria was higher in patients with diverticulitis than in controls, and patients clustered separately from controls based on Proteobacteria profiles. Kvasnovsky examined 30 patients with SUDD and compared the 15 with a prior history of diverticulitis to those without prior diverticulitis. A history of diverticulitis was associated with higher relative abundance of *Pseudobutyrivibrio, Bifidobacterium*, Christensenellaceae family and Mollicutes RF9.(25)

Our study extends the findings of these prior studies in several important ways. First, we used next-generation, high-throughput DNA sequencing technology to enable deep, quantitative profiling of the fecal microbiota in diverticulitis and diverticulosis. This methodology captures a comprehensive distribution of organisms that would be missed with fingerprinting approaches such as denaturing gradient gel electrophoresis (DGGE) as were used in 2 of the 3 prior studies of diverticulitis. Indeed, the differences we found between cases and controls were predominantly in less abundant organisms. Second, we selected comparable controls with asymptomatic diverticulosis matched on age and sex in an attempt to elucidate a microbial profile that may indicate or predispose to disease progression from diverticulosis to diverticulitis. In comparison, prior studies used unmatched controls with colon cancer, IBD and irritable bowel syndrome (IBS) (i.e. diseases associated with specific dysbioses), who were not known to have diverticulosis, or studied SUDD, a different entity from diverticulitis. In addition, controls in the study of Daniels et al (29) had undergone a colon preparation immediately prior to sample collection. Bowel preparation is known to significantly disrupt the intestinal microbiota.(57) For example, in a study by Jalanka et al, immediately following polyethylene glycol preparation, the total microbial load was decreased 31-fold and nearly one fourth of patients lost the participant-specificity of their microbiota.(58) Therefore, it is likely that the differences reported by Daniels et al reflect at least in part the colon purge given to the controls but not the diverticulitis cases. Third, we analyzed the fecal microbiome as a representation of the entire intestinal bacterial community. In contrast, in a prior study of diverticulitis (30) the microbiome was sampled via rectal swabs.(29) The mucosal-associated microbiota of the rectum are known to differ from the microbiota in the large and small intestine and represent a specific microbial niche.(59) Lastly, we collected data on important potential confounders of the intestinal microbial composition including diet, BMI and smoking that were not accounted for in prior studies.

There are several potential mechanisms by which the fecal microbiota may contribute to the development of diverticulitis. A long-held theory suggests that stasis within diverticula leads to local bacterial overgrowth and inflammation. Our results suggest that a dysbiosis in the intestine in general, and not just within a specific diverticulum, might also predispose to or precipitate diverticulitis potentially through pro-inflammatory effects or impaired gut barrier function.(60, 61) In a global analysis of the microbiome, we found no association between the clusters of highly abundant bacteria (Bacteroides, Ruminococcus, and Prevotella) and diverticulitis; however, we detected differences in more minor components of the microbiome suggesting that the development of diverticulitis may be influenced by alteration in these less-abundant groups. In particular, we found higher levels of Akkermansia, Marvinbyrantia, Anaerotruncus, Subdolignulum, and Coriobacteria in patients with diverticulitis compared to those with diverticulosis. Several of these genera have been associated with inflammatory conditions of the intestine. (62-68) Succinate-producing genera such as Marvinbryantia have been shown to promote *C. difficile* infection after antibiotic treatment, (69) suggesting that alterations in the metabolic capacity of the microbiome could be important for the development of diverticulitis. Lastly, Subdoligranulum has been associated with increased secretion of the proinflammatory cytokine IL-β in patients with post-infectious IBS.(67) On the other hand, we found a decrease in two genera associated with short chain fatty acid production, Coprococcus and Barnesiella, in diverticulitis cases vs controls (although fiber intake was higher on average in cases than in controls). Short chain fatty acid production by the microbiome is generally associated with intestinal health. Coprococcus has been found in reduced numbers in patients with IBS (70) and IBD,(68) whereas the expansion of Barnesiella upon dietary oligosaccharide feeding has been shown to protect against colitis in mice (71).

Of interest, we found a higher proportion of cases were of Hispanic ethnicity than controls (40% vs 0%). This may reflect the demographic at one of the participating hospitals, the willingness of certain ethnicities to participate, a higher prevalence of certain risk factors in this group, or an underlying difference in the risk of diverticulitis. Several, small studies report a high proportion of Hispanic males, especially those with obesity, among young patients with diverticulitis.(72, 73) Future studies should explore ethnic differences in the risk of diverticulitis.

Our study has several strengths. As noted above, we selected age and gender-matched controls with asymptomatic diverticulosis seen at colonoscopy, collected data on a variety of factors that might confound the relationship between the intestinal microbiota and diverticulitis, including diet and other lifestyle variables, and used next-generation sequencing techniques to enable deeper investigation of the fecal microbial community than prior studies that generally used fingerprinting approaches. In addition, we recruited prospective cases with CT-proven diverticulitis in an effort to capture the microbial communities that might predispose to the development of this condition.

Diverticulitis is acute in onset, and in the U.S. it is treated promptly with antibiotics. These factors complicate the recruitment of study participants and the collection of baseline or pre-diverticulitis fecal samples. We identified patients presenting to the hospital with acute diverticulitis via daily searches of radiology and emergency department logs as well as a radiology alert system. However, most patients with diverticulitis are diagnosed in the outpatient setting making it difficult to identify and recruit cases prior to the receipt of antibiotics. Even in the inpatient setting, an additional 6 patients were initially recruited for the study, but became ineligible due to the use of antibiotics at the time of recruitment or receipt of antibiotics or surgery between the collection of the first and second fecal sample. We collected the first stool after presentation in an attempt to capture the baseline microbial communities prior to antibiotics. However, all patients had received some antibiotics prior to stool collection (mean 1 day). This brief antibiotic exposure may have influenced the bacterial communities in addition to the localized inflammation associated with diverticulitis. In addition, the lower abundance of bacterial groups such as *Coriobacteria, Erysipeolotrichaceae* and *Akkermansia* and increase in others such as *Proteobacter* and *Verruncomicrobia* may reflect a response to antibiotics.(74) Therefore, we also collected fecal samples at least two months following treatment. However, microbial communities may not yet have returned to baseline. We found that the alpha-diversity was lower in post-treatment cases than in diverticulitis cases at diagnosis although the diversity was more similar to controls post-treatment. Overall, it is unclear whether the differences seen between cases and controls are the cause or result of diverticulitis or treatment.

Our sample size was small, limiting our ability to differentiate cases from controls. Many of our findings were of borderline significance or lost significance when adjusted for multiple testing. In addition, our ability to account for and balance important confounders in cases vs controls was limited. For example, fiber intake was significantly higher in cases than controls. It is possible that cases were instructed to increase fiber intake. However, only 2 (20%) of cases had a prior diagnosis of diverticulitis or diverticulosis. In comparison, all controls had a previous diagnosis of diverticulosis. Furthermore, fiber intake was very similar at the time of diagnosis and two months later in cases. Nonetheless, it is possible that knowledge of the relationship between fiber and diverticular disease impacted dietary recall. We adjusted for fiber intake in our multivariable models and found no significant association between fiber and the overall microbial composition of dominant organisms.

To overcome some of the difficulties associated with the analysis of the fecal microbiome in acute diverticulitis, we are expanding our work to include a study of patients with a history of diverticulitis. A prospective study of patients with diverticulosis would be the optimal means to study the role of the gut microbiome in the development of diverticulitis; however, given that fewer than 5% of patients with diverticulosis develop diverticulitis over an average of 11 years of follow up,(32) such a study would need to be large and have long-term follow-up.

In conclusion, our findings indicate that the composition of the fecal microbiota in patients with diverticulitis differs from that of matched controls with asymptomatic diverticulosis. Defining such differences in the fecal microbiota has important potential clinical implications since they could be modeled to enable outcome prediction and risk stratification, provide avenues for diagnosis, treatment, and prevention, and reveal insight into disease pathogenesis.

## Data Availability

Sequences have been deposited in the Sequence Read Archive of NCBI under accession number SUB2127005

## References

1. Peery AF, Crockett SD, Barritt AS, Dellon ES, Eluri S, Gangarosa LM, et al. Burden of Gastrointestinal, Liver, and Pancreatic Diseases in the United States. Gastroenterology. 2015;149(7):1731–41 e3.

2. Peery AF, Dellon ES, Lund J, Crockett SD, McGowan CE, Bulsiewicz WJ, et al. Burden of gastrointestinal disease in the United States: 2012 update. Gastroenterology. 2012;143(5):1179–87 e3.

3. Peery AF, Crockett SD, Murphy CC, Lund JL, Dellon ES, Williams JL, et al. Burden and Cost of Gastrointestinal, Liver, and Pancreatic Diseases in the United States: Update 2018. Gastroenterology. 2019;156(1):254–72 e11.

4. Berman LG, Burdick D, Heitzman ER, Prior JT. A critical reappraisal of sigmoid peridiverticulitis. Surg Gynecol Obstet. 1968;127(3):481–91.

5. Floch MH, White J. Diverticulitis: new concepts and new therapies. J Clin Gastroenterol. 2005;39(5):355–6.

6. Strate LL, Modi R, Cohen E, Spiegel BM. Diverticular disease as a chronic illness: evolving epidemiologic and clinical insights. Am J Gastroenterol. 2012;107(10):1486–93.

7. Strate LL, Morris AM. Epidemiology, Pathophysiology, and Treatment of Diverticulitis. Gastroenterology. 2019;156(5):1282–98 e1.

8. Feingold D, Steele SR, Lee S, Kaiser A, Boushey R, Buie WD, et al. Practice parameters for the treatment of sigmoid diverticulitis. Dis Colon Rectum. 2014;57(3):284–94.

9. Cao Y, Strate LL, Keeley BR, Tam I, Wu K, Giovannucci EL, et al. Meat intake and risk of diverticulitis among men. Gut. 2017.

10. Strate LL, Keeley BR, Cao Y, Wu K, Giovannucci EL, Chan AT. Western Dietary Pattern Increases, and Prudent Dietary Pattern Decreases, Risk of Incident Diverticulitis in a Prospective Cohort Study. Gastroenterology. 2017;152(5):1023–30 e2.

11. Crowe FL, Balkwill A, Cairns BJ, Appleby PN, Green J, Reeves GK, et al. Source of dietary fibre and diverticular disease incidence: a prospective study of UK women. Gut. 2014;63(9):1450–6.

12. Crowe FL, Appleby PN, Allen NE, Key TJ. Diet and risk of diverticular disease in Oxford cohort of European Prospective Investigation into Cancer and Nutrition (EPIC): prospective study of British vegetarians and non-vegetarians. BMJ. 2011;343:d4131.

13. Ma W, Jovani M, Liu PH, Nguyen LH, Cao Y, Tam I, et al. Association Between Obesity and Weight Change and Risk of Diverticulitis in Women. Gastroenterology. 2018.

14. Strate LL, Liu YL, Aldoori WH, Syngal S, Giovannucci EL. Obesity increases the risks of diverticulitis and diverticular bleeding. Gastroenterology. 2009;136(1):115–22 e1.

15. Louis P, Scott KP, Duncan SH, Flint HJ. Understanding the effects of diet on bacterial metabolism in the large intestine. J Appl Microbiol. 2007;102(5):1197–208.

16. Turnbaugh PJ, Gordon JI. The core gut microbiome, energy balance, and obesity. J Physiol. 2009.

17. Martinez-Medina M, Denizot J, Dreux N, Robin F, Billard E, Bonnet R, et al. Western diet induces dysbiosis with increased E coli in CEABAC10 mice, alters host barrier function favouring AIEC colonisation. Gut. 2014;63(1):116–24.

18. Claesson MJ, Jeffery IB, Conde S, Power SE, O’Connor EM, Cusack S, et al. Gut microbiota composition correlates with diet and health in the elderly. jature. 2012;488(7410):178–84.

19. Cotillard A, Kennedy SP, Kong LC, Prifti E, Pons N, Le Chatelier E, et al. Dietary intervention impact on gut microbial gene richness. Nature. 2013;500(7464):585–8.

20. De Filippis F, Pellegrini N, Vannini L, Jeffery IB, La Storia A, Laghi L, et al. High-level adherence to a Mediterranean diet beneficially impacts the gut microbiota and associated metabolome. Gut. 2016;65(11):1812–21.

21. De Filippo C, Cavalieri D, Di Paola M, Ramazzotti M, Poullet JB, Massart S, et al. Impact of diet in shaping gut microbiota revealed by a comparative study in children from Europe and rural Africa. Proc Natl Acad Sci U S A. 2010;107(33):14691–6.

22. Tomkins AM, Bradley AK, Oswald S, Drasar BS. Diet and the faecal microflora of infants, children and adults in rural Nigeria and urban U.K. J Hyg (Lond). 1981;86(3):285–93.

23. Kang JY, Dhar A, Pollok R, Leicester RJ, Benson MJ, Kumar D, et al. Diverticular disease of the colon: ethnic differences in frequency. Aliment Pharmacol Ther. 2004;19(7):765–9.

24. Barbara G, Scaioli E, Barbaro MR, Biagi E, Laghi L, Cremon C, et al. Gut microbiota, metabolome and immune signatures in patients with uncomplicated diverticular disease. Gut. 2017;66(7):1252–61.

25. Kvasnovsky CL, Leong LEX, Choo JM, Abell GCJ, Papagrigoriadis S, Bruce KD, et al. Clinical and symptom scores are significantly correlated with fecal microbiota features in patients with symptomatic uncomplicated diverticular disease: a pilot study. European journal of gastroenterologyhepatology. 2018;30(1):107–12.

26. Tursi A, Mastromarino P, Capobianco D, Elisei W, Miccheli A, Capuani G, et al. Assessment of Fecal Microbiota and Fecal Metabolome in Symptomatic Uncomplicated Diverticular Disease of the Colon. J Clin Gastroenterol. 2016;50 Suppl 1:S9–S12.

27. Baugh MD, Perry MJ, Hollander AP, Davies DR, Cross SS, Lobo AJ, et al. Matrix metalloproteinase levels are elevated in inflammatory bowel disease. Gastroenterology. 1999;117(4):814–22.

28. Jones RB, Fodor AA, Peery AF, Tsilimigras MCB, Winglee K, McCoy A, et al. An Aberrant Microbiota is not Strongly Associated with Incidental Colonic Diverticulosis. Scientific reports. 2018;8(1):4951.

29. Daniels L, Budding AE, de Korte N, Eck A, Bogaards JA, Stockmann HB, et al. Fecal microbiome analysis as a diagnostic test for diverticulitis. European journal of clinical microbiologyinfectious diseases : official publication of the European Society of Clinical Microbiology. 2014;33(11):1927–36.

30. Gueimonde M, Ouwehand A, Huhtinen H, Salminen E, Salminen S. Qualitative and quantitative analyses of the bifidobacterial microbiota in the colonic mucosa of patients with colorectal cancer, diverticulitis and inflammatory bowel disease. World J Gastroenterol. 2007;13(29):3985–9.

31. Peery AF, Keku TO, Martin CF, Eluri S, Runge T, Galanko JA, et al. Distribution and Characteristics of Colonic Diverticula in a United States Screening Population. Clin Gastroenterol Hepatol. 2016;14(7):980–5 e1.

32. Shahedi K, Fuller G, Bolus R, Cohen E, Vu M, Shah R, et al. Long-term risk of acute diverticulitis among patients with incidental diverticulosis found during colonoscopy. Clin Gastroenterol Hepatol. 2013;11(12):1609–13.

33. Strate LL, Peery AF, Neumann I. American Gastroenterological Association Institute Technical Review on the Management of Acute Diverticulitis. Gastroenterology. 2015;149(7):1950–76 e12.

34. White E, Patterson RE, Kristal AR, Thornquist M, King I, Shattuck AL, et al. VITamins And Lifestyle cohort study: study design and characteristics of supplement users. Am J Epidemiol. 2004;159(1):83–93.

35. Li F, Hullar MA, Lampe JW. Optimization of terminal restriction fragment polymorphism (TRFLP) analysis of human gut microbiota. J Microbiol Methods. 2007;68(2):302–11.

36. Dowd SE, Sun Y, Wolcott RD, Domingo A, Carroll JA. Bacterial tag-encoded FLX amplicon pyrosequencing (bTEFAP) for microbiome studies: bacterial diversity in the ileum of newly weaned Salmonella-infected pigs. Foodborne Pathog Dis. 2008;5(4):459–72.

37. Schloss PD. A high-throughput DNA sequence aligner for microbial ecology studies. PLoS One. 2009;4(12):e8230.

38. Schloss PD, Westcott SL, Ryabin T, Hall JR, Hartmann M, Hollister EB, et al. Introducing mothur: Open-Source, platform-independent, community-supported software for describing and comparing microbial communities. Appl Environ Microbiol. 2009;75(23):7537–41.

39. Edgar RC, Haas BJ, Clemente JC, Quince C, Knight R. UCHIME improves sensitivity and speed of chimera detection. Bioinformatics. 2011;27(16):2194–200.

40. Quince C, Lanzen A, Davenport RJ, Turnbaugh PJ. Removing Noise From Pyrosequenced Amplicons. BMC Bioinformatics. 2011;12:DOI: 10.1186/471-2105-12-38.

41. Schloss PD. Secondary structure improves OTU assignments of 16S rRNA gene sequences. ISME J. 2013;7(3):457–60.

42. Yilmaz P, Parfrey LW, Yarza P, Gerken J, Pruesse E, Quast C, et al. The SILVA and “All-species Living Tree Project (LTP)” taxonomic frameworks. Nucleic Acids Res. 2014;42(Database issue):D643–8.

43. Pruesse E, Quast C, Knittel K, Fuchs BM, Ludwig WG, Peplies J, et al. SILVA: a comprehensive online resource for quality checked and aligned ribosomal RNA sequence data compatible with ARB. Nucleic Acids Res. 2007;35(21):7188–96.

44. Huse SM, Welch DM, Morrison HG, Sogin ML. Ironing out the wrinkles in the rare biosphere through improved OTU clustering. Environ Microbiol. 2010;12(7):1889–98.

45. Claesson MJ, O’Toole PW. Evaluating the latest high-throughput molecular techniques for the exploration of microbial gut communities. Gut Microbes. 2010;1(4):277–8.

46. Navas-Molina JA, Peralta-Sanchez JM, Gonzalez A, McMurdie PJ, Vazquez-Baeza Y, Xu Z, et al. Advancing our understanding of the human microbiome using QIIME. Methods Enzymol. 2013;531:371–444.

47. Lin JH. Divergence measures based on the Shannon Entropy. IEEE Trans Info Theory. 1991;37(1):145–51.

48. Arumugam M, Raes J, Pelletier E, Le Paslier D, Yamada T, Mende DR, et al. Enterotypes of the human gut microbiome. Nature. 2011;473(7346):174–80.

49. McCune B, Grace JB. Analysis of ecological communities. 1 ed. Glendenen Beach, OR: MjM Software; 2002. 284 p.

50. Benjamini Y, Hochberg Y. Controlling the False Discovery Rate - a Practical and Powerful Approach to Multiple Testing. Journal of the Royal Statistical Society Series B-Methodological. 1995;57(1):289–300.

51. Worley B, Powers R. Mulitvariate Analysis in Metabolomics. Current Metabolomics. 2013;1:92–107.

52. Westerhuis JA, Hoefsloot HCJ, Smit S, Vis DJ, Smilde AK, van Velzen EJJ, et al. Assessment of PLSDA cross validation. Metabolomics. 2008;4(1):81–9.

53. Worley B, Powers R. Multivariate Analysis in Metabolomics. Current Metabolomics. 2013;1(1):92–107.

54. Mehmood TL, KH; Snipen, L; Saebo, S. A review of variable selection methods in Partial Least Squares Regression. Chemometr Intell Lab. 2012;118:62–9.

55. Magurran A. Diversity indices and species abundance models. Ecological Diversity and Its Measurement. Princeton, NJ: Princeton University Press; 1988. p. 8-45.

56. Arumugam M, Raes J, Pelletier E, Le Paslier D, Yamada T, Mende DR, et al. Enterotypes of the human gut microbiome. Nature. 2011;473(7346):174–80.

57. Drago L, Toscano M, De Grandi R, Casini V, Pace F. Persisting changes of intestinal microbiota after bowel lavage and colonoscopy. European journal of gastroenterologyhepatology. 2016;28(5):532–7.

58. Jalanka J, Salonen A, Salojarvi J, Ritari J, Immonen O, Marciani L, et al. Effects of bowel cleansing on the intestinal microbiota. Gut. 2015;64(10):1562–8.

59. Li G, Yang M, Zhou K, Zhang L, Tian L, Lv S, et al. Diversity of Duodenal and Rectal Microbiota in Biopsy Tissues and Luminal Contents in Healthy Volunteers. Journal of microbiology and biotechnology. 2015;25(7):1136–45.

60. Duncan SH, Holtrop G, Lobley GE, Calder AG, Stewart CS, Flint HJ. Contribution of acetate to butyrate formation by human faecal bacteria. Br J Nutr. 2004;91(6):915–23.

61. Pryde SE, Duncan SH, Hold GL, Stewart CS, Flint HJ. The microbiology of butyrate formation in the human colon. FEMS Microbiol Lett. 2002;217(2):133–9.

62. Baxter NT, Zackular JP, Chen GY, Schloss PD. Structure of the gut microbiome following colonization with human feces determines colonic tumor burden. Microbiome. 2014;2:20.

63. Zackular JP, Baxter NT, Iverson KD, Sadler WD, Petrosino JF, Chen GY, et al. The gut microbiome modulates colon tumorigenesis. mBio. 2013;4(6):e00692–13.

64. Weir TL, Manter DK, Sheflin AM, Barnett BA, Heuberger AL, Ryan EP. Stool microbiome and metabolome differences between colorectal cancer patients and healthy adults. PLoS One. 2013;8(8):DOI 10.1371/journal.pone.0070803.

65. Ijssennagger N, Belzer C, Hooiveld GJ, Dekker J, van Mil SWC, Muller M, et al. Gut microbiota facilitates dietary heme-induced epithelial hyperproliferation by opening the mucus barrier in colon. Proc Natl Acad Sci U S A. 2015;112(32):10038–43.

66. Morgan XC, Kabakchiev B, Waldron L, Tyler AD, Tickle TL, Milgrom R, et al. Associations between host gene expression, the mucosal microbiome, and clinical outcome in the pelvic pouch of patients with inflammatory bowel disease. Genome Biol. 2015;16(1):67.

67. Sundin J, Rangel I, Repsilber D, Brummer RJ. Cytokine Response after Stimulation with Key Commensal Bacteria Differ in Post-Infectious Irritable Bowel Syndrome (PI-IBS) Patients Compared to Healthy Controls. Plos One. 2015;10(9).

68. Shaw KA, Bertha M, Hofmekler T, Chopra P, Vatanen T, Srivatsa A, et al. Dysbiosis, inflammation, and response to treatment: a longitudinal study of pediatric subjects with newly diagnosed inflammatory bowel disease. Genome medicine. 2016;8(1):75.

69. Passalacqua KD, Charbonneau ME, O’Riordan MX. Bacterial Metabolism Shapes the Host-Pathogen Interface. Microbiol Spectr. 2016;4(3).

70. Kassinen A, Krogius-Kurikka L, Makivuokko H, Rinttila T, Paulin L, Corander J, et al. The fecal microbiota of irritable bowel syndrome patients differs significantly from that of healthy subjects. Gastroenterology. 2007;133(1):24–33.

71. Weiss GA, Chassard C, Hennet T. Selective proliferation of intestinal Barnesiella under fucosyllactose supplementation in mice. Br J Nutr. 2014;111(9):1602–10.

72. Schauer PR, Ramos R, Ghiatas AA, Sirinek KR. Virulent diverticular disease in young obese men. American journal of surgery. 1992;164(5):443–6; discussion 6-8.

73. Syed U, Companioni R, Bansal R, Alkhawam H, Walfish A. Diverticulitis in the Young Population: Reconsidering Conventional Recommendations. Acta gastro-enterologica Belgica. 2016;79(2):435–9.

74. Dubourg G, Lagier JC, Armougom F, Robert C, Audoly G, Papazian L, et al. High-level colonisation of the human gut by Verrucomicrobia following broad-spectrum antibiotic treatment. International journal of antimicrobial agents. 2013;41(2):149–55.

